# DeepACSA: Automatic segmentation of anatomical cross-sectional area in ultrasound images of human lower limb muscles using deep learning

**DOI:** 10.1101/2021.12.27.21268258

**Authors:** Paul Ritsche, Philipp Wirth, Neil J. Cronin, Fabio Sarto, Marco V. Narici, Oliver Faude, Martino V. Franchi

**Affiliations:** Department of Sport, Exercise & Health, University of Basel, Switzerland; Lightly AG, Zurich, Switzerland; Neuromuscular Research Centre, Faculty of Sport and Health Sciences, University of Jyvaskyla, Finland; Department of Biomedical Sciences, University of Padova, Italy

**Author notes:** These authors share last authorship (Oliver Faude, Martino V. Franchi). Corresponding author: Paul Ritsche, Department of Sport, Exercise & Health, University of Basel, Grosse Allee 6, 4052 Basel.

**Keywords:** deep learning, image segmentation, muscle, ultrasound, U-net

## Abstract

**Background:** Muscle anatomical cross-sectional area (ACSA) is an important parameter that characterizes muscle function and helps to classify the severity of several muscular disorders. Ultrasound is a patient friendly, fast and cheap method of assessing muscle ACSA, but manual analysis of the images is laborious, subjective and requires thorough experience. To date, no open access and fully automated program to segment ACSA in ultrasound images is available. On this basis, we present DeepACSA, a deep learning approach to automatically segment ACSA in panoramic ultrasound images of the human rectus femoris (RF), vastus lateralis (VL), gastrocnemius medialis (GM) and lateralis (GL) muscles.

**Methods:** We trained convolutional neural networks using 1772 ultrasound images from 153 participants (25 females, 128 males; mean age = 38.2 years, range: 13-78) captured by three experienced operators using three distinct devices. We trained three muscle-specific models to detect ACSA.

**Findings:** Comparing DeepACSA analysis of the RF to manual analysis resulted in intra-class correlation (ICC) of 0.96 (95% CI 0.94,0.97), mean difference of 0.31 cm^2^ (0.04,0.58) and standard error of the differences (SEM) of 0.91 cm^2^ (0.47,1.36). For the VL, ICC was 0.94 (0.91,0.96), mean difference was 0.25 cm^2^ (−0.21,0.7) and SEM was 1.55 cm^2^ (1.13,1.96). The GM/GL muscles demonstrated an ICC of 0.97 (0.95,0.98), a mean difference of 0.01 cm^2^ (−0.25, 0.24) and a SEM of 0.69 cm^2^ (0.52,0.83).

**Interpretation:** DeepACSA provides fast and objective segmentation of lower limb panoramic ultrasound images comparable to manual segmentation and is easy to implement both in research and clinical settings. Inaccurate model predictions occurred predominantly on low-quality images, highlighting the importance of high image quality for accurate prediction.

**Research in context:** *Evidence before this study:* Lower limb muscle cross-sectional area is an important predictor of physical performance, frailty, and it can be used in the diagnosis of sarcopenia or in the monitoring of several muscular disorders. Panoramic ultrasound has been proven valid in obtaining images of human muscles compared to magnetic resonance imaging. Further, ultrasound can be performed on bedside and in patients unable to undergo Magnetic Resonance Imaging, in example intensive care unit patients. However, post-scanning manual segmentation of muscle cross-sectional area is laborious and subjective. Thus, automatization of the segmentation process would benefit both researchers and clinicians. We searched Pubmed from database inception to August 31, 2021, using the search terms “deep learning” OR “machine learning” AND “ultrasound” AND “muscle” AND “cross sectional area”. The search yielded 15 results, with two investigations comparing deep learning based analysis of lower limb muscle cross-sectional area ultrasound images to manual evaluation. By using the bibliographies of the retrieved articles, we identified another investigation. However, none of the found investigations included panoramic ultrasound images displaying a whole muscle cross-sectional area in their data sets.

*Added value of this study:* We developed DeepACSA, an open-source tool to automatically segment the anatomical cross-sectional area in ultrasound images of human lower limb muscles. This is, to our knowledge, the first deep learning based algorithm segmenting panoramic ultrasound images. In contrast to previously proposed algorithms, we used panoramic ultrasound images. DeepACSA analysis was comparable to manual segmentation and reduced time of analysis. Thus, the value added by this investigation lies in increased efficiency and reduced subjectivity of muscle cross-sectional area segmentation. DeepACSA includes a graphical user interface allowing for straight forward implementation.

*Implications of all the available evidence:* Incorporating deep learning based algorithms which automate the segmentation of muscle cross-sectional area in clinical practice can reduce analysis effort and operator bias. DeepACSA can be easily implemented in clinical and research settings, allowing for fast evaluation of large image datasets. Research is ongoing to assess the generalizability of our results in ageing and pathological settings, and to other imaging modalities.

## Introduction

The anatomical cross-sectional area of a muscle (ACSA) represents a two-dimensional index of muscle size acquired in the transversal plane.^1^ Muscle ACSA is important for clinical and scientific practice; it is related to the capacity of a muscle to generate force and, as a consequence, to locomotor performance.^2,3^ Recent investigations have shown that muscle ACSA represents a useful parameter in the diagnosis and classification of several muscular disorders as well as the potential monitoring of disease progression.^4–7^ Lower limb muscle ACSA is often assessed in the diagnosis of sarcopenia and is strongly associated with frailty.^4–6^ Moreover, measuring lower limb muscle mass or ACSA is necessary to assess the extent of induced muscle loss in cachectic, dystrophic, and even intensive care unit patients.^7–9^ Muscle mass is also a predictor of hospital stay duration in patients with moderate to severe COVID-19.^10^ Thus, whole muscle ACSA represents a crucial variable when monitoring muscle decline in pathological settings, in response to disuse and ageing, and when investigating muscle adaptations to training in rehabilitation and return to sport scenarios.

Muscle ACSA can be assessed using several techniques such as Magnetic Resonance Imaging (MRI) and computer tomography, or with ultrasound imaging. Due to technical advances, the ability to perform measurements at the bedside, and drastically lower costs compared to other imaging modalities, the use of ultrasound has increased in many research and clinical settings.^4,11,12^ Because of the shape and size of several lower limb muscles, conventional static B-mode ultrasound is often unsuitable to assess whole muscle ACSA.^1^ The main reason for this is the limited field of view of most transducers.^1,11^ Hence, panoramic ultrasound, which is used to obtain a panoramic image of the whole muscle, is employed to circumvent the limitations imposed by the field of view of most commercially available transducers^1^. In fact, panoramic ultrasound has previously shown good comparability to MRI and excellent inter-session and inter-rater repeatability.^13,14^

Once ultrasound is implemented as a routine part of ACSA monitoring, large volumes of image data could be collected, necessitating efficient and reliable analysis methods. To date, ultrasound images of muscle ACSA are mostly evaluated manually. However, manual analysis is subjective, laborious, and requires great experience.^15,16^ Furthermore, manually analysing large image datasets is not only time consuming but also leads to reduced focus and thus error proneness.^16^ Semi-automated and automated algorithms have been developed to accelerate the ACSA segmentation process and reduce the subjectivity of manual segmentation in lower limb muscle ultrasound images.^17,18^ These approaches used sophisticated image processing steps to localize features and objects within an image. However, because of this, their generalisability is limited, and they present inconsistent results depending on image properties.

Making use of deep learning might be advantageous compared to non-trainable image processing alone, especially since convolutional neural networks (CNNs) have shown great promise identifying features in medical ultrasound and MRI scans.^12,16^ U-net structured CNNs^19^ have inter alia been successfully applied to brain MRI segmentation^20^, carotid plaque segmentation in ultrasound images^21^, as well as lower limb muscle segmentation in ultrasound and MRI images.^22–25^ Previously, Chen et al.^24^ and Marzola et al.^22,23^ presented automatic approaches to segment the ACSA of upper and lower limb muscles, but only in conventional static brightness mode ultrasound images, and not panoramic images.

To our knowledge, the semi-automated ACSAuto program^17^ is the only open-access tool available for analysing panoramic lower limb ultrasound images. Thus, it should be determined whether CNN approaches could be used to fully automate the process of ACSA analysis from panoramic ultrasound images. This could increase objectivity and reduce the time and effort needed for the analysis process.^15,22^ Although manual ACSA segmentation of ultrasound images is the current gold standard, using automatic approaches might increase the evaluation quality.

In this study we present DeepACSA, a python package incorporating U-net based CNNs to automatically segment the ACSA of lower limb muscles in panoramic ultrasound images. DeepACSA is open source and includes a custom graphical user interface (GUI) to allow straightforward use and implementation. The muscles of interest are the m. rectus femoris (RF), m. vastus lateralis (VL), mm. gastrocnemius medialis (GM) and lateralis (GL). Here we describe the DeepACSA package and compare it to manual image analysis and a non-trainable image processing script (ACSAuto)^17^.

## Methods

### Image acquisition and data

The data used in this study contains images from 143 participants of different age groups (adolescent males: n = 50, 16·4 years (13 to 17), adults: n = 83 (25 females, 58 males), 26·9 years (18 to 40), elderly males: n=10, 71·5 years (65 to 78)). We used 602 ultrasound images of the RF, 634 ultrasound images of the VL and 298 ultrasound images of the GM and GL (9 to 36 images per participant) acquired in previous and ongoing studies which received ethical approval from the responsible committees. Images were anonymized and randomly divided into a training set for the models as well as an external validation set, making sure that no image of the same muscle region of one participant appeared in both sets. To increase image variability, we used brightness mode panoramic ultrasonography from three devices (ACUSON Juniper, linear-array 54 mm probe, 12L3, Acuson 12L3, SIEMENS Healthineers, Erlangen, Germany; Aixplorer Ultimate, linear-array 38mm probe, Superline SL10-2, SuperSonic Imagine, Aix-en-Provence, France; Mylab 70, linear array 47mm probe, Esaote Biomedica, Genova, Italy) collected by three different experienced operators to assess the ACSA of RF, VL, GM and GL. We acquired scans at rest while the participants laid in a supine position with their legs extended and feet on the bed. A guide was mounted to the leg to control the transversal path. Because of regional differences in muscle size and shape, we acquired images of the RF and VL at 10% increments from 30 to 70% of the distance between the lateral femur condyle and the trochanter major. We acquired images of the GM and GL at 30 and 50% of muscle length. Scans of the RF and VL muscles were either taken separately or cropped from whole quadriceps scans to increase image variability. Ultrasound device settings differed between devices and study protocols, and image depth, brightness and contrast settings were always chosen to ensure best visibility of the muscle. Images included in the training and test sets were manually labelled by an experienced investigator (PR) prior to model training (Fig. 1). Manual segmentation consisted of digitizing the ACSA of each muscle using the polygon tool in FIJI^26^. We randomly included 30 images from the proximal (70% of femur length), mid (50% of femur length) and distal (30% of femur length) regions in the validation set for the RF and VL. For the GM and GL, 37 images from the mid (50% of muscle length) and 28 images from the proximal (30% of muscle length) region were randomly included in the validation set. Therefore, the validation set consisted of 90 images for the RF and VL and 65 images for the GM and GL. The ratio of images in the validation and training sets for the RF was 0·18 (90:512), for the VL 0·17 (90:544), and for the GM and GL 0·27 (65:233). No cropped images were included in the validation set because we only had these images available from one device. We measured ACSA in all images of the validation set using the DeepACSA and ACSAuto programs. Because GL segmentation is not supported in ACSAuto, we only analyzed the GM validation images with ACSAuto.

**Figure 1:**
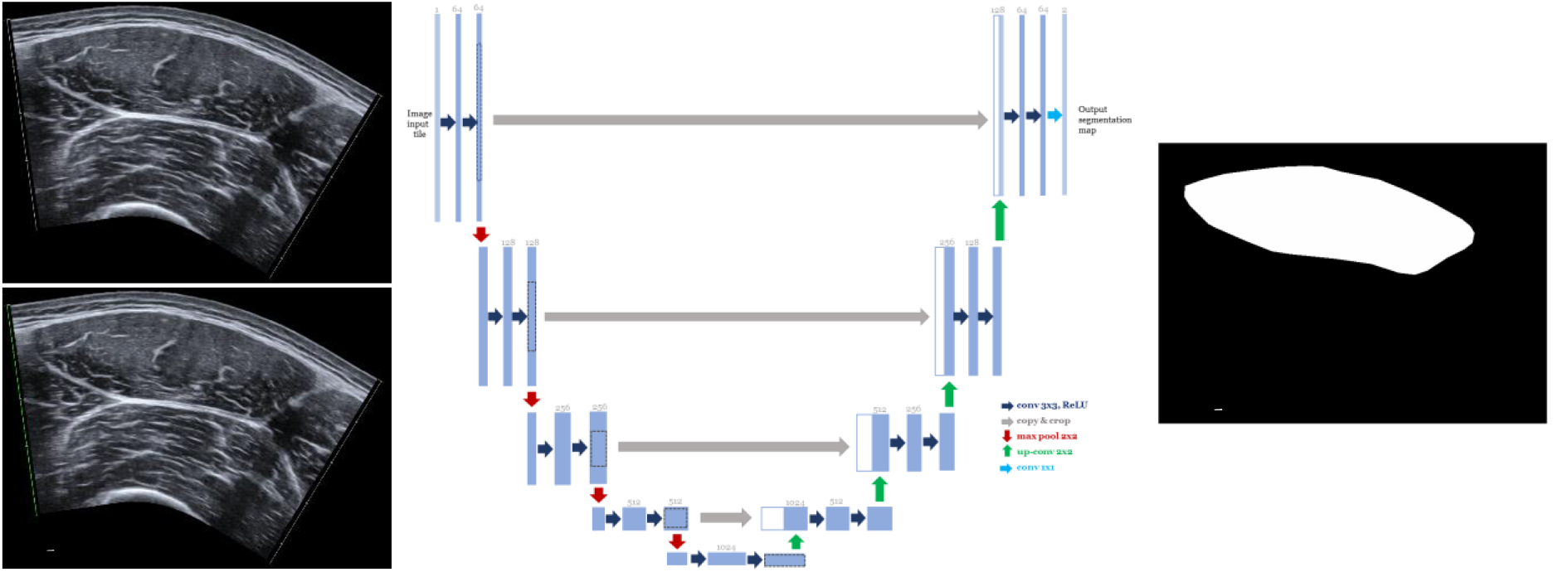
DeepACSA workflow. a) Original ultrasound image of the m. rectus femoris (RF) at 50% of femur length with automatic scaling (green line). b) The original image is then preprocessed with contrast-limited adaptive histogram equalization and inputted to the model. c) Detailed U-net CNN architecture (modified from Ronneberger et al.21 and Cronin et al.15). Multi-channel feature maps are represented by the blue boxes with number of channels displayed on top of the respective box. Copied feature maps from the convolutional (left) side that are concatenated with the ones from the expanding (right) side are represented by the white boxes. The different operations are marked by the arrows. d) Model prediction of muscle area following post-processing (shown as a binary image).

### Network architecture and training

We performed sematic segmentation with two classes (ACSA and background) on pre-processed ultrasound images. The image-label pairs (Fig. 1) served as input for the models. The models we employed have a U-net architecture (Fig. 2), consisting of a contracting and an expanding path.^15,19^ The exact architecture of the U-net model can be found elsewhere.^15,19^ The encoder (left) path uses convolution and max pooling layers to generate a 1024-dimensional abstract representation of the input image. Subsequently, the decoder (right) path uses deconvolutional layers to generate a pixelwise class predicton from the abstract image representaion of the input. Features between layers of the same resolution are shared (Fig. 2) so that information about small-scale objects is not lost (hence the U-shape). The model outputs a pixelwise binary label, thus, every pixel of an image is predicted to belong to one of two possible classes. We adopted U-net models due to their demonstrated success for image segmentation in several fields^15,22–25^ and their ability to work well with small datasets containing a few hundred images^19^. Prior to model training, images and masks in the training sets of all muscles were augmented using height and width shift, rotation and horizontal flipping (see shared code for details). We imported, normalized to a scale between 0 and 1, and resized the images to 256 × 256 pixels for training. Training the CNNs with 512 × 512-pixel sized images yielded inferior results and was computationally more expensive. Images were preprocessed with contrast-limited adaptive histogram equalization to increase contrast between aponeurosis and muscle tissue. By convention, we applied a random 90/10 % training/test data split within the training data set.^15^ We trained three separate models, one for RF, one for VL and one for GM and GL combined with the same U-net model architecture.^15,19^ A RTX3060 GPU (NVIDIA, Santa Clara, USA) was used for model training with a defined maximum of 50 epochs and a batch size of one. We used the Adam optimizer for stochastic gradient descent to update network weights with an initial learning rate of 10^−5^.^15,19^ The learning rate was reduced by a factor of 0.1 following ten epochs of loss stagnation. We used binary cross-entropy as a loss function because the segmentation task only represents two classes. To reduce the risk of overfitting, we implemented early stopping when the training error decreased, and the test error reached a plateau or increased. During training, model performance was evaluated using the intersection over union (IoU) measure to test the overlap between manually segmented muscle area and labels predicted by the respective model (Table 1). IoU is defined as

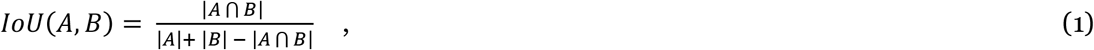

where 0 ≤ IoU (A, B) ≤ 1, A describes the ground truth and B the model predictions. The IoU is maximal if the ground truth A and model predictions B are the same and minimal if A and B have no overlap.

**Figure 2:**
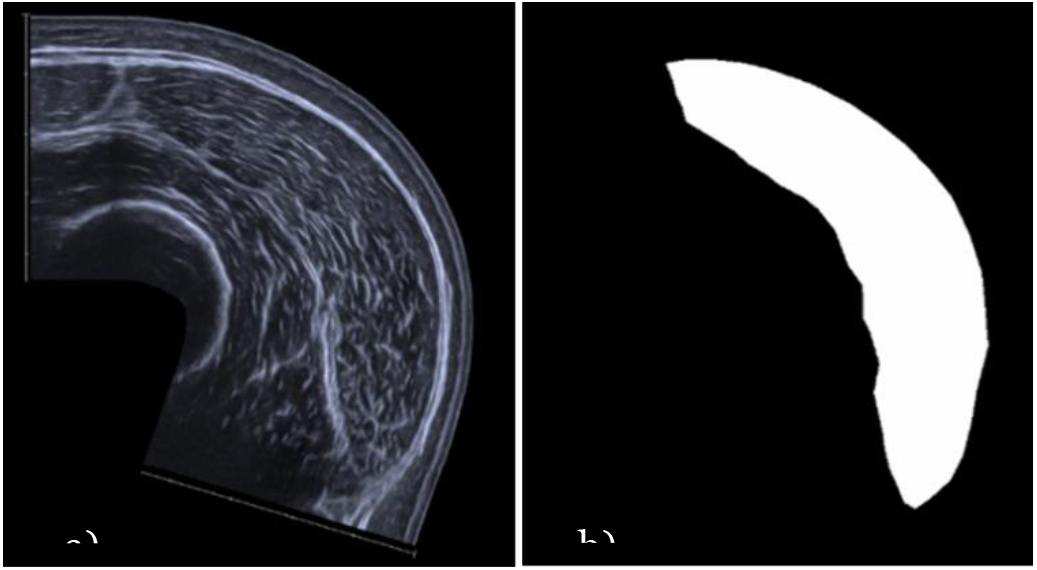
Manual segmentation for convolutional neural network training. a) Original extended-field-of-view ultrasound image of m. vastus lateralis, b) manually segmented binary mask of muscle in a).

**Table 1.** Final intersection over union and loss functions for the three selected models.

| Metric | RF |  | VL |  | GM & GL |  |
| --- | --- | --- | --- | --- | --- | --- |
|  | Training | Test | Training | Test | Training | Test |
| IoU | 0.997 | 0.996 | 0.994 | 0.992 | 0.997 | 0.996 |
| Accuracy | 0.986 | 0.984 | 0.975 | 0.971 | 0.975 | 0.973 |

Final intersection over union (IoU) and accuracy metrics for the training and test sets. IoU was calculated for the model prediction and the manually segmented ground truth in the training and the randomly generated test set using the training/test split. *M. rectus femoris* (RF), *m. vastus lateralis* (VL), *mm. gastrocnemius medialis* (GM), and *lateralis* (GL).

The output of each CNN is a binary mask image that contains the predicted muscle area. The output is post-processed so that holes within the predicted muscle are filled and, in case of multiple predicted structures, only the largest structure is kept. The DeepACSA code is written in Python using a Keras API to interface with Tensorflow for model training. The code and trained models from this project are openly available at https://github.com/PaulRitsche/DeepACSA.git. For more information about program use we refer the reader to an instructional online video (Link: https://youtu.be/It9CqVSNc9M).

### Analysis metrics

All statistical analyses were performed in R software ^27^ (Base, BlandAltmanLeh, irr, readxl and rstudioapi packages). The analysis script is shared as a supplementary file. We compared DeepACSA and ACSAuto measurements to manual measurements. For this purpose, we calculated consecutive-pairwise intra-class correlations (ICC) and standard error of the differences between methods (SEM) with 95% compatibility intervals (CI). We used Bland–Altman analysis^28^ to test the agreement between two analysis methods and set the limits of agreement to ± 1.96 standard deviations. We computed the standardized mean bias according to Hopkins^29^, with 0·1, 0·3, 0·6, 1·0 and 2·0 representing small, moderate, large, very large and extremely large errors respectively.

## Results

The training of the deep neural networks required 23 to 38 epochs, and IoU and loss metrics are shown in Table 1. An example of pre-processing and model prediction following post-processing is shown in Figure 2. ICCs, SEMs, mean bias, standardized mean bias with 95% CI as well as percentage values for all muscles comparing DeepACSA and ACSAuto to manual measurements are shown in Table 2 and Fig. 3. We additionally report comparability statistics after removing incorrect predictions (RF: 5·5%, VL: 13·3%, GM & GL: 3·1%) by the models (Table 2). Predictions were removed following visual inspections by an experienced investigator (PR) based on obvious deviations from the expected ACSA of the respective muscle (Fig. 4). Comparing DeepACSA analysis of all muscles including the wrong predictions to manual analysis resulted in ICCs between 0·94 and 0·99, mean differences of -0·01 to 0·37 cm^2^ and SEMs ranging from 0·34 to 1·55 cm^2^ (Table 2). Calculated standardized mean biases were small for all muscles ranging from -0·02 to 0·07 (Table 2). The comparison of ACSAuto with manual analysis of all images and muscles resulted in ICCs between 0·97 and 0·99, mean differences of -0·13 to 0·24 cm^2^ and SEMs ranging from 0·23 to 0·66 cm^2^. Standardized mean biases were small for all muscles ranging from -0·02 to 0·05 (Table 2). DeepACSA segmentation resulted in differences between muscle regions for the RF. At 30% of femur length, the ICC was 0·503 (95% CI: 0·18;0·73), the mean difference was 0·79 cm^2^ (0·03;1·55) and the SEM was 1.44 cm^2^ (0·69;2·12). Standardized mean bias was moderate (0·40). In contrast, at 50% of femur length, the ICC was 0·91 (0·83;0·96), the mean difference was 0·07 cm^2^ (−0·21;0·36) and the SEM was 0·54 cm^2^ (0·18;0·86). Standardized mean bias was small (0·04). At 70% of femur length, the ICC was 0·97 (0·94;0·99), the mean difference was 0·06 cm^2^ (−0·03;0·15) and the SEM was 0.18 cm^2^ (0.11;0.23). Similar to the 50% of femur length site, the observed standardized mean bias was small (0·05). Regional data for VL, GM & GL can be found in the supplementary material. Removing incorrect model predictions resulted in increased ICCs and decreased mean differences, SEMs, and standardized mean bias for all muscles (Table 2). In Bland-Altman analysis, concerns regarding heteroscedasticity were only raised when comparing manual to DeepACSA RF analysis. With increasing muscle size, it seems that the variance around the mean difference increases.

**Table 2.**
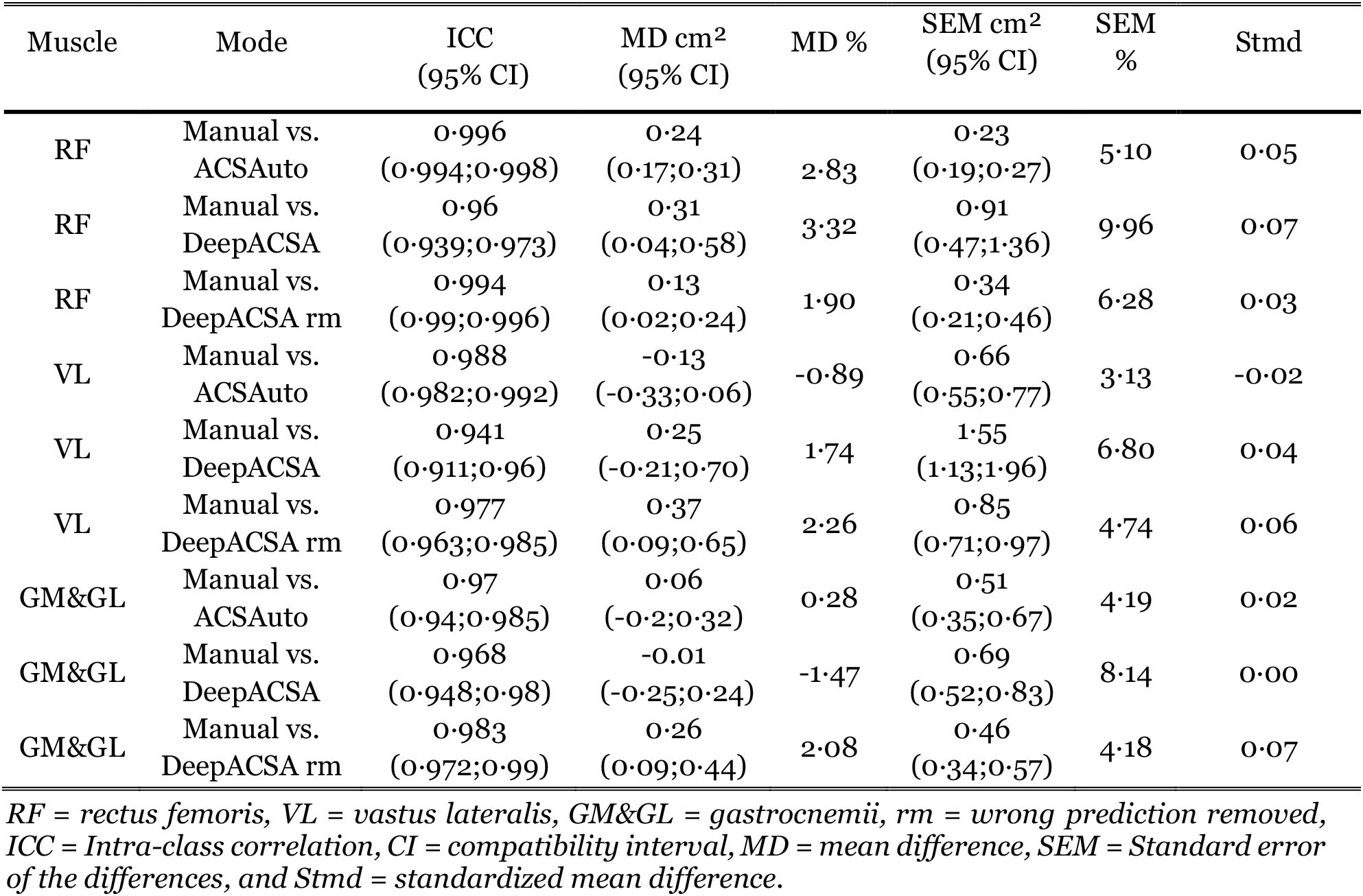
Comparison of manual analysis and ACSAuto and DeepACSA

**Figure 3:**
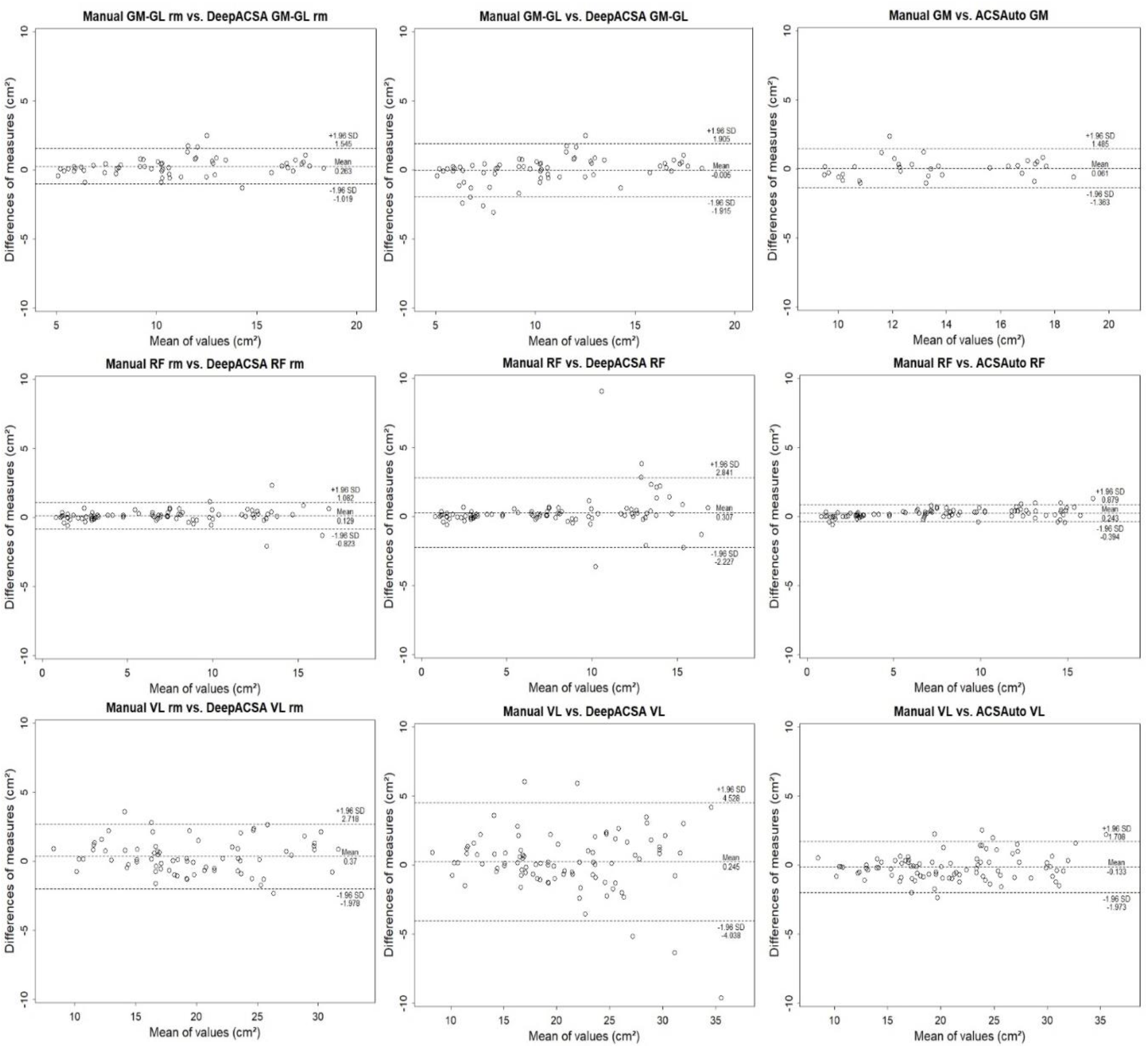
Bland-Altman plots of all muscles plotting the difference between manual and DeepACSA with incorrect predictions removed (rm), manual and DeepACSA as well as manual and ACSAuto area segmentation measurements against the mean of both measures. Dotted and solid lines illustrate 95% limits of agreement and bias. M. rectus femoris (RF) and m. vastus lateralis (VL), mm. gastrocnemius medialis (GM), and lateralis (GL).

**Figure 4:**
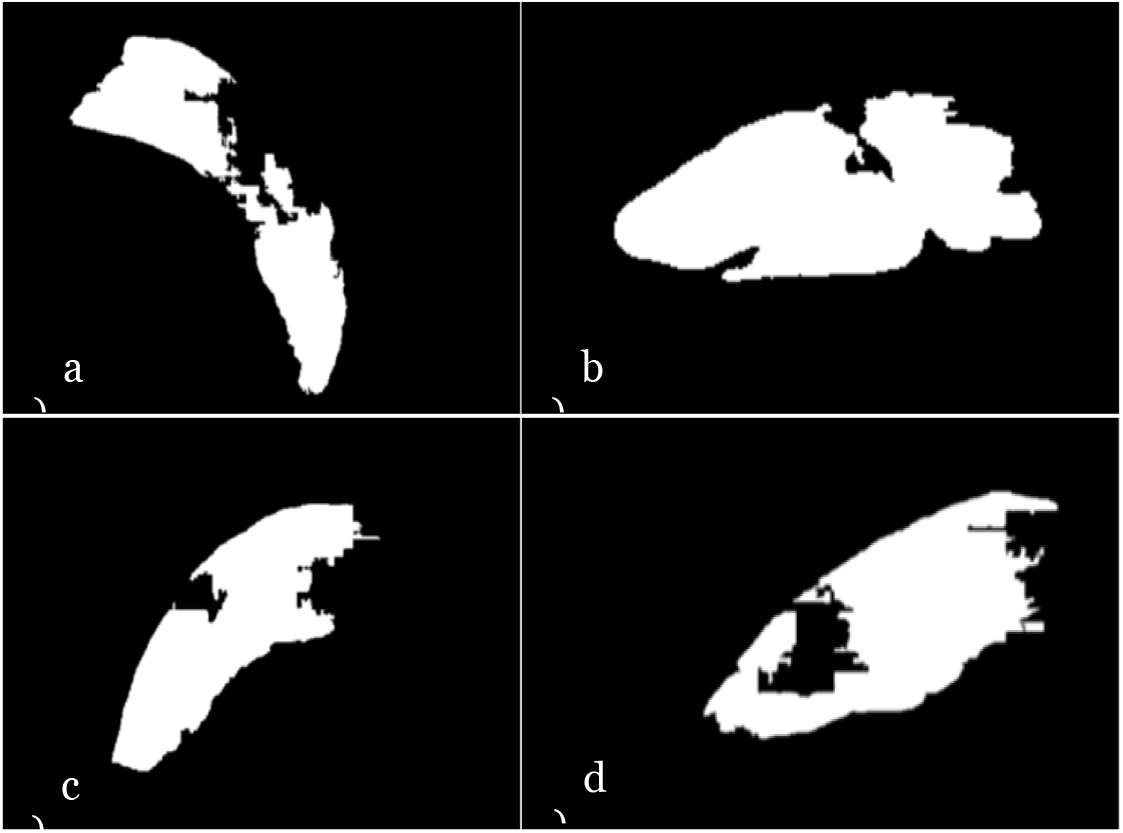
Examples of incorrect predictions and obvious deviations from the expected ACSA of the respective muscles. a) m. vastus lateralis, b) m. rectus femoris, c) m. gastrocnemius medialis, and d) m. gastrocnemius lateralis.

Additionally, we compared the efficiency of DeepACSA and ACSAuto. Whereas analysis duration was 4.0s standard deviation (SD) ± 0.43 for analyzing one image in our validation set usind DeepACSA, analysis duration for one image using ACSAuto was 40.3s ± SD 6.9.

## Discussion

Here we present DeepACSA, a deep learning approach for automatic analysis of muscle ACSA in panoramic brightness mode ultrasound images. The predictions of our trained CNNs were comparable to manual segmentation for all the lower limb muscles investigated. To our knowledge, DeepACSA represents the first openly accessible program able to automatically segment the muscle area of rectus femoris, vastus lateralis, gastrocnemius medialis, and gastrocnemius lateralis in panoramic ultrasound images.

Comparing the DeepACSA-based analysis of all muscles, the predictions of GM & GL ACSA were found to have the highest agreement with manual analysis. The worst agreement was observed for the RF, which was also the only muscle that displayed regional differences. Most incorrect RF predictions occurred at 30% of femur length. Here the aponeuroses of the RF are more difficult to scan, and thus the image quality decreases. More distally, at 70% of femur length, the RF is usually small, sometimes non-existent, and the echogenicity is increased, leading to reduced overall pixel contrast. Small muscle area and lower pixel contrast make prediction more difficult and might lead to deletion or selection of the wrong area during post-processing.

All our CNNs demonstrated minimal loss and high IoU scores. Although high IoU and minimal loss values in the training set might indicate overfitting, similar values were observed for the test set. Thus, our model can generalize to unseen data. However, we used ultrasound images of all three devices in our training, test and validation sets leading to similar image characteristics in all data sets. To test whether our CNNs can generalize to unseen image data from other devices, images from a fourth device should solely be added to the validation set. Based on our results, visual inspection of the model output is still necessary, and the removal of erroneous predictions resulted in increased comparability for all muscles. However, we believe that adding more training data might help to eliminate these model failures. Erroneous predictions in the validation set mostly occurred when image quality was low, i.e. when tissue contrast was low or pixel values were homogeneous. Therefore, image quality is still an important factor for correct muscle ACSA segmentation using deep learning. Images for which muscle ACSA was predicted incorrectly should be analyzed using ASCAuto^17^ or manually.

Given that DeepACSA is comparable to manual evaluation, the program might be implementable in a clinical setting as well. In contrast to MRI or computer tomography, ultrasound can be used quickly at the bedside of patients and practitioners do not necessarily need active cooperation from patients.^4^ This might be especially important for patients in the intensive care unit or with severe muscular disorders.^9,30^ DeepACSA analysis is comparable to manual evaluation, more time efficient, and automates the segmentation process. This is important for clinical practice as it decreases the effort needed to analyze ultrasound image data. In fact, reducing the subjectivity of the analysis might increase the overall validity of clinical results. Moreover, our trained CNNs could be used to correctly predict the ACSA of other muscles, in patients with muscular diseases or an ageing population (for example when screening for the presence of sarcopenia). However, because of differences in image properties, it might be necessary to retrain our models including labeled images from the respective muscles or clinical populations in the training dataset. In the future, we aim to release more models that allow data from additional muscles and clinical populations to be analysed using the DeepACSA package.

In contrast to the algorithms presented by Chen et al.^24^ and Marzola et al.^22,23^, who used a supervised learning approach with U-net structured CNNs and achieved good precision and recall rates, DeepACSA can segment muscles in panoramic ultrasound images. To our knowledge, DeepACSA is the only program that includes images of several lower limb muscles from various muscle regions, different operators, and several ultrasound devices. Whereas Marzola et al.^22^ included images from the tibialis anterior, GM and biceps brachii at rest, Chen et al.^24^ included images of the RF during contraction. Acquiring images during contraction could limit the generalizability of the model to images acquired at rest. Furthermore, while the algorithms proposed by Marzola et al.^22,23^ seem to distinguish between clinical and healthy populations based on echogenicity values, segmenting ACSA in muscles that exceed the field of view of the ultrasound transducer may have limited meaningfulness.

It is important to note that the predictions of our trained CNNs for all muscles showed lower agreement with manual analysis when compared to ACSAuto (DeepACSA: 95% limits of agreement 4·5 to -4·0 cm^2^, ACSAuto: 1·7 to -1·9 cm^2^). This is due to the manual correction of the suggested outline during ACSAuto analysis. Yet, the feature detection filters used for image segmentation in ACSAuto are highly dependent on image properties. Furthermore, ACSAuto is somewhat subjective because the user must validate the proposed muscle outline. In addition, not correcting the suggested outline led to large measurement errors compared to manual analysis and thus unusable results.^17^ By employing trainable CNNs, DeepACSA is more robust to variation in ultrasound image pixel characteristics because the whole image texture is considered, and more complex features are computed. Additionally, DeepACSA reduces user bias in the analysis process, as no user input is required during image segmentation.

This investigation has some limitations. First, we performed no cross-validation or hyper parameter tuning for our models.^16^ Additionally, comparing several different deep neural networks with various architectures could be beneficial to determine the ideal model for this task. However, the U-net architecture we employed demonstrated good results when segmenting muscle aponeuroses and fascicles in sagittal plane ultrasound images of the VL and GM.^15^ Although our training set consisted of data from different age groups, we only included images from adolescent and young healthy participants and three devices in our validation sets because of limited available data from elderly people. Thus, we cannot yet generalize our results to older people, clinical populations or other devices.^22,23^ Finally, DeepACSA is currently able to automatically evaluate single muscle images but not videos.

## Conclusion

DeepACSA segmentation of panoramic ultrasound images from rectus femoris, vastus lateralis, gastrocnemius medialis and lateralis muscles yielded comparable results to manual segmentation. Therefore, our trained convolutional neural networks can automatically segment lower limb muscles in panoramic ultrasound images. DeepACSA objectifies and accelerates the evaluation process of panoramic ACSA ultrasound images, allowing large datasets to be evaluated quickly, and representing a valuable tool that can be implemented in clinical settings. However, our results demonstrated that visual inspection of the output may still be necessary to optimize predictions and avoid misclassifications. In the future, the segmentation performance of different model architectures should be compared whilst using a larger, more variable training dataset.

## Supporting information

Supplementary Table 1. Regional ACSA segmentation for VL and GM / GL.

R analysis script.

## Data Availability

All data used in this study is publicly available. The DeepACSA package is available on Github at https://github.com/PaulRitsche/DeepACSA.git. Our trained models are available at https://doi.org/10.5281/zenodo.5799068. Anonymized panoramic ultrasound images are available at https://doi.org/10.5281/zenodo.5799204.

https://doi.org/10.5281/zenodo.5799068

https://doi.org/10.5281/zenodo.5799204

## Data sharing statement

All data used in this study is publicly available. The DeepACSA package is available on Github at https://github.com/PaulRitsche/DeepACSA.git. Our trained models are available at https://doi.org/10.5281/zenodo.5799068. Anonymized panoramic ultrasound images are available at https://doi.org/10.5281/zenodo.5799204. The instructional video can be accessed using this link https://youtu.be/It9CqVSNc9M.

## Contributors

P.R., O.F., and M.V.F designed the study. M.V.N. and N.J.C. assisted in designing the study. P.R., P.W., and N.J.C. wrote the DeepACSA package code. P.R., F.S., and M.V.F. collected the ultrasound images. P.R. and M.V.F. performed annotations and quality control. P.R. and P.W. performed the deep learning analysis and visualizations. P.R., O.F. and M.V.F wrote the first draft of the manuscript. All authors read and revised the manuscript draft and approved the final manuscript. All authors had access to all the data in the study and P.R., O.F. and M.V.F verified the data and had final responsibility for the decision to submit for publication.

